# The spectrum of psychiatric manifestations in subacute sclerosing panencephalitis: a systematic review of published case reports and case series

**DOI:** 10.1101/2023.05.15.23290012

**Authors:** Ravindra Kumar Garg, Sujita Kumar Kar, Hardeep Singh Malhotra, Shweta Pandey, Amita Jain, Imran Rizvi, Ravi Uniyal, Neeraj Kumar

## Abstract

**Background:** Data related to psychiatric manifestations in SSPE is currently available only in form of isolated case reports. In this systematic review, we evaluated the spectrum of psychiatric manifestations and their impact on the course and outcome of SSPE.

**Methods:** Data were obtained from four databases (PubMed, Embase, Scopus, and Google Scholar), with the most recent search conducted on 27/03/2023. The PRISMA guidelines were followed, and the PROSPERO registration number for the protocol is CRD42023408227. SSPE was diagnosed using Dyken’s criteria. Extracted data were recorded in an Excel spreadsheet. To evaluate the quality of the data, the Joanna Briggs Institute Critical Appraisal tool was employed.

**Results:** Our search resulted in 30 published reports of 32 patients. The mean age was 17.9 years. Schizophrenia, catatonia, and poorly characterized psychotic illnesses were the three most common psychiatric presentations (60%) in SSPE cases. Mania or depression was reported among 23% (7/32) cases. In 10% of cases, the initial clinical presentation of SSPE was considered functional/ hysterical. In approximately 81% (26/32) cases, the course of SSPE was rapidly progressive (either acute fulminant or subacute). Treatment with antipsychotic drugs had poor or no response. Out of 17 patients who received antipsychotic drugs 6 patients noted severe extrapyramidal adverse effects, that further deteriorated the clinical condition of the patients.

**Conclusion:** Several patients with SSPE inadvertently end up in psychiatric care due to some psychiatric manifestation. Early psychiatric disorders in SSPE are often subtle and diagnosis of SSPE is easily missed.

## Introduction

Subacute sclerosing panencephalitis (SSPE) is a relentlessly progressive brain disorder caused by the defective measles virus. SSPE is common in measles-endemic areas. SSPE usually presents with progressive cognitive deterioration and myoclonus. Children between 8 and 12 years of age are more frequently affected. SSPE is invariably fatal within 1–3 years from the onset. In an acute-fulminant variant of SSPE, the patient either dies or becomes akinetic mute within 6 months of disease onset. Electroencephalography (EEG) reveals periodic discharges. Brain imaging reveals periventricular T2/ FLAIR white matter abnormalities. The advanced stage of SSPE is characterized by marked brain atrophy. A definitive diagnosis needs the demonstration of elevated measles antibody titers in the cerebrospinal fluid (CSF).**^1,2^** Infrequently, pure psychiatric manifestations (like mania, psychosis, catatonia, and malingering) have also been reported in many SSPE patients. In this systematic review, we evaluated the spectrum of psychiatric manifestations of SSPE.

## Material and methods

### Protocol registration

We conducted a thorough analysis of published case reports and case series of SSPE patients, who either had psychiatric manifestations at the time of diagnosis or who later developed psychiatric manifestations. We used the Preferred Reporting Items for Systematic Reviews and Meta-Analyses (PRISMA) standards for our systematic review. The protocol was made pre-registered in PROSPERO (CRD42023408227).**^3^**

### Literature search

A literature search was done in the databases of PubMed, Scopus, Embase, and Google Scholar. In Google Scholar database, first 50 pages were screened for relevant articles. No language restrictions were imposed, while databases are searched. Articles, other than in English language were translated into English with the help of “A Google translator”.

The following search items were used in our search strategy: (((((((((((((PSYCHIATRIC) OR (Neuropsychiatric)) OR (Schizophrenia)) OR (Mania)) OR (Psychosis)) OR (Depression)) OR (Catatonia)) OR (Attention deficits)) OR (Anxiety)) OR (Panic attacks)) OR (Sleep disorders)) OR (Nightmares)) OR (Parasomnias)) AND (SSPE). The date of the last search was 27/03/2023.

### Exclusion criteria

The articles that were omitted, included editorials, comments on previously published cases, and review articles. Conference abstracts were also not taken into account.

### Data extraction

The study was conducted in two stages. Titles and abstracts were assessed by two independent reviewers in the initial phase (RK and IR). The full texts of the chosen papers were then examined by an additional set of two reviewers to determine their eligibility following the inclusion criteria (RK and IR). A third author was brought in to settle any differences between the two authors (AJ).

Dyken’s criteria were used to identify cases with a confirmed diagnosis of SSPE (Table-1).**^4^**Studies were considered if they fulfilled the following criteria: (1) they were case reports or case series; (2) they described cases of psychiatric manifestations in a case of SSPE; (3) cohort studies were included only if, individual patient data were available; and (4) they comprehensively described the psychiatric manifestations in confirmed SSPE cases.

**Table-1:**
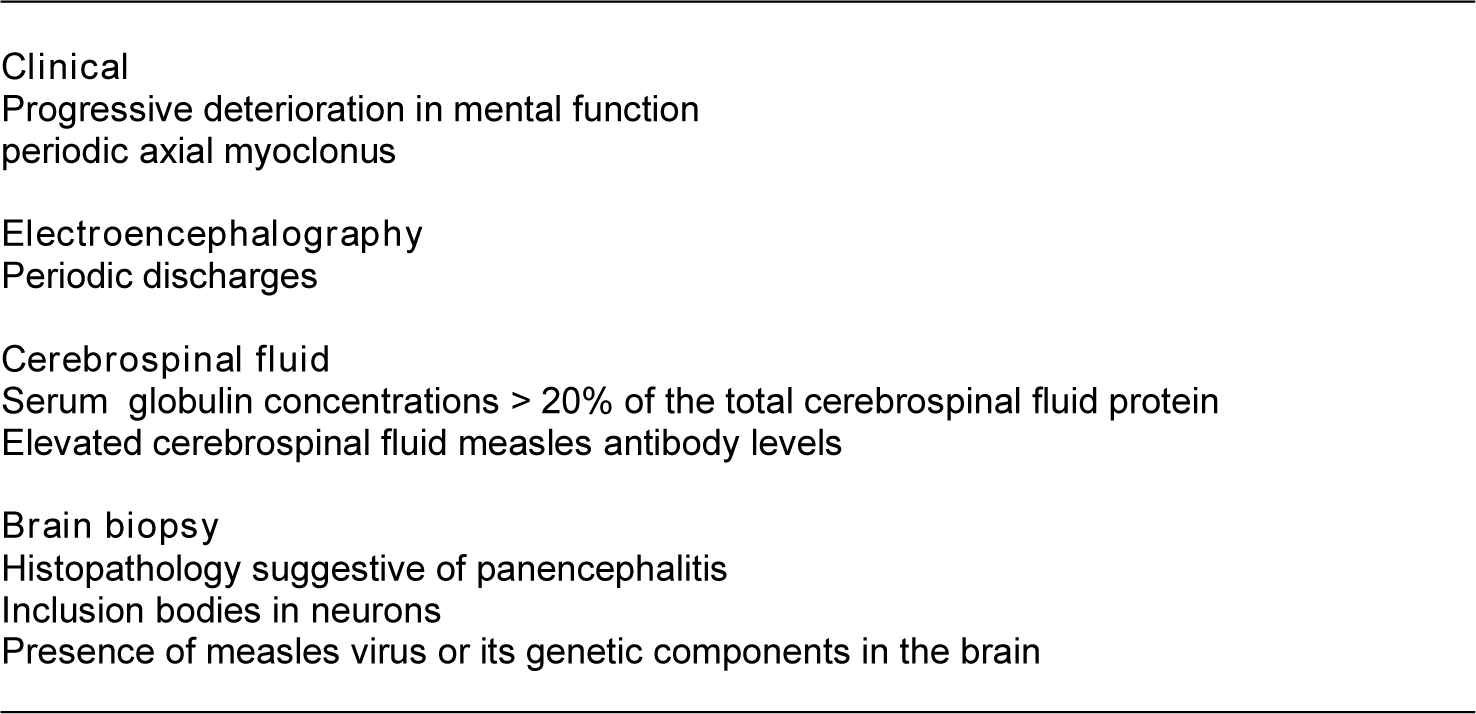
Dyken’s diagnostic criteria of subacute sclerosing panencephalitis (SSPE) (Any three of 4 components)

### Quality of studies

The critical appraisal checklist for case reports and case series given by the Joanna Briggs Institute (JBI) was used to assess the quality of published case reports.

The critical appraisal tool we used was an 8-item scale including the patient’s demographic characteristics, clinical details, details of laboratory workup, the treatment administered, the follow-up clinical condition, adverse events, and the main takeaways from the case reports. Each of these criteria was used to evaluate every case. “Yes” or “No” was indicated for each point.**^5^** Two independent reviewers (SKK and RU) assessed the quality of the included cases, and any disagreements between them were resolved by mutual agreement, If still there was a dispute, it was resolved via discussion with a third reviewer (AJ).

### Data analysis

Each patient’s demographic information, history of measles infection or vaccination in infancy, length of illness, type of psychiatric manifestations, workup procedures, results of neuroimaging, and course and outcome were all noted. Four reviewers (RK, IR, HSM, and RU) completed all of them; in the event of a disagreement, assistance from a fifth reviewer was sought (AJ). To manage duplicate records, the web tool EndNote 20 (Clarivate Analytics) was employed. Two reviewers independently completed this process once more (SKK and AJ). Any problems were rectified with the assistance of another reviewer. A PRISMA flow chart was used to present the number of records that were retrieved and evaluated at each stage. PRISMA flow chart was prepared with the help of EndNote 20 (Clarivate Analytics). (Figure-1)

**Figure-1.**
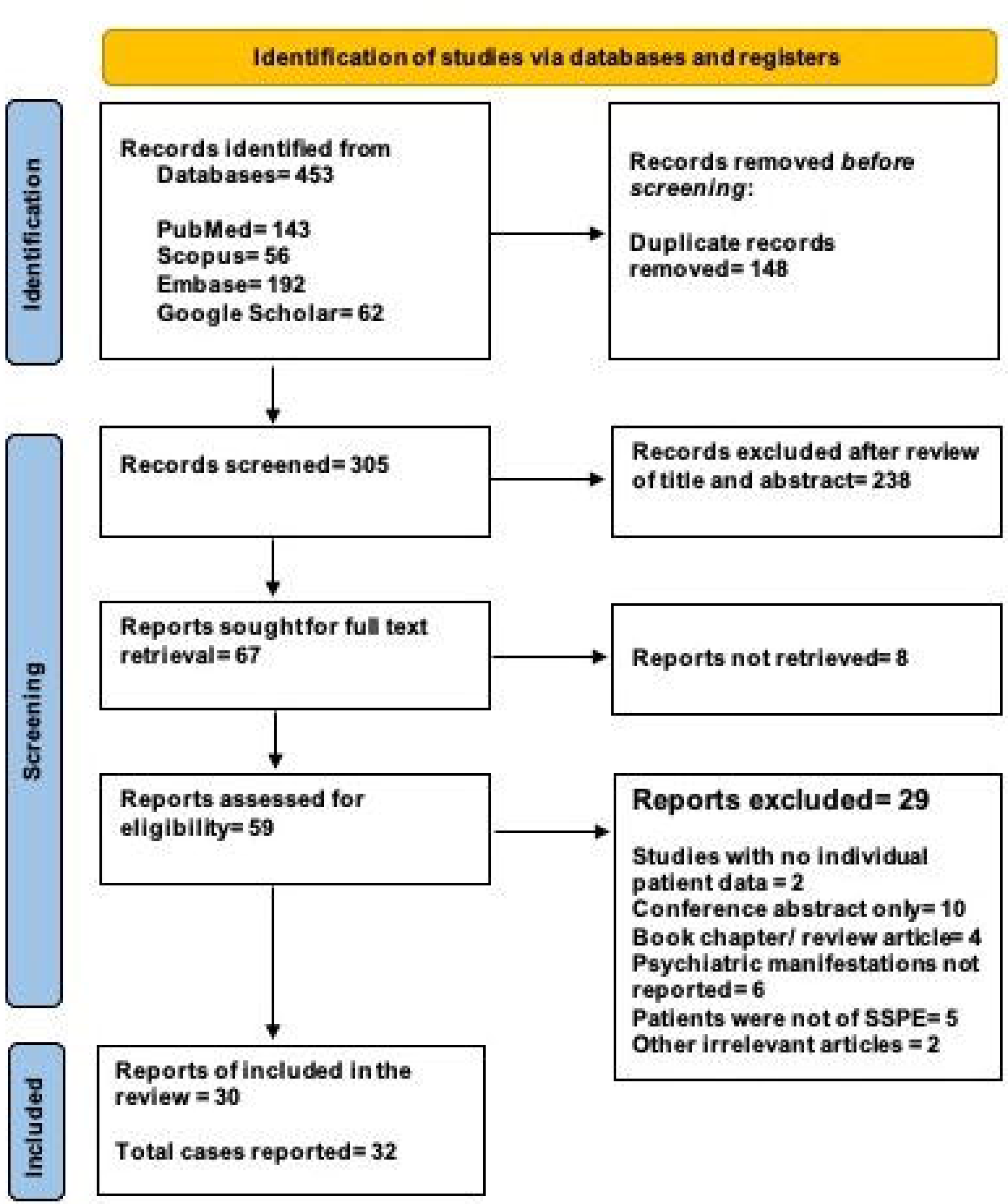
PRISMA flow diagram of the study shows the process of article selection for systematic review.

We used Microsoft Excel software, for data analysis. The necessary data was taken out and put together in an Excel document. The information we compiled included the first author, the nation of the report, the demographics of the patients, the status of childhood measles vaccination, childhood measles infection, length of illness, types of psychiatric manifestations, other clinical presentations, specifics of the reported diagnostic workup, neuroimaging, and the outcome. We focussed on the descriptive and qualitative aspects of the data. Frequencies and percentages were employed to represent the categorical variables. Means, medians, or ranges were employed for reporting continuous variables.

## Results

Our search resulted in 30 reports with information on 32 patients. (Table-2)**^6-35^** We compiled our data as per PRISMA standards. (Supplementary file-1-PRISMA checklist) Figure-1 shows the PRISMA flowchart for our systematic review. Table-3 summarises the demographic, clinical, neuroimaging, and brain/retina biopsy information of SSPE cases with psychiatric manifestations.

**Table-2:**
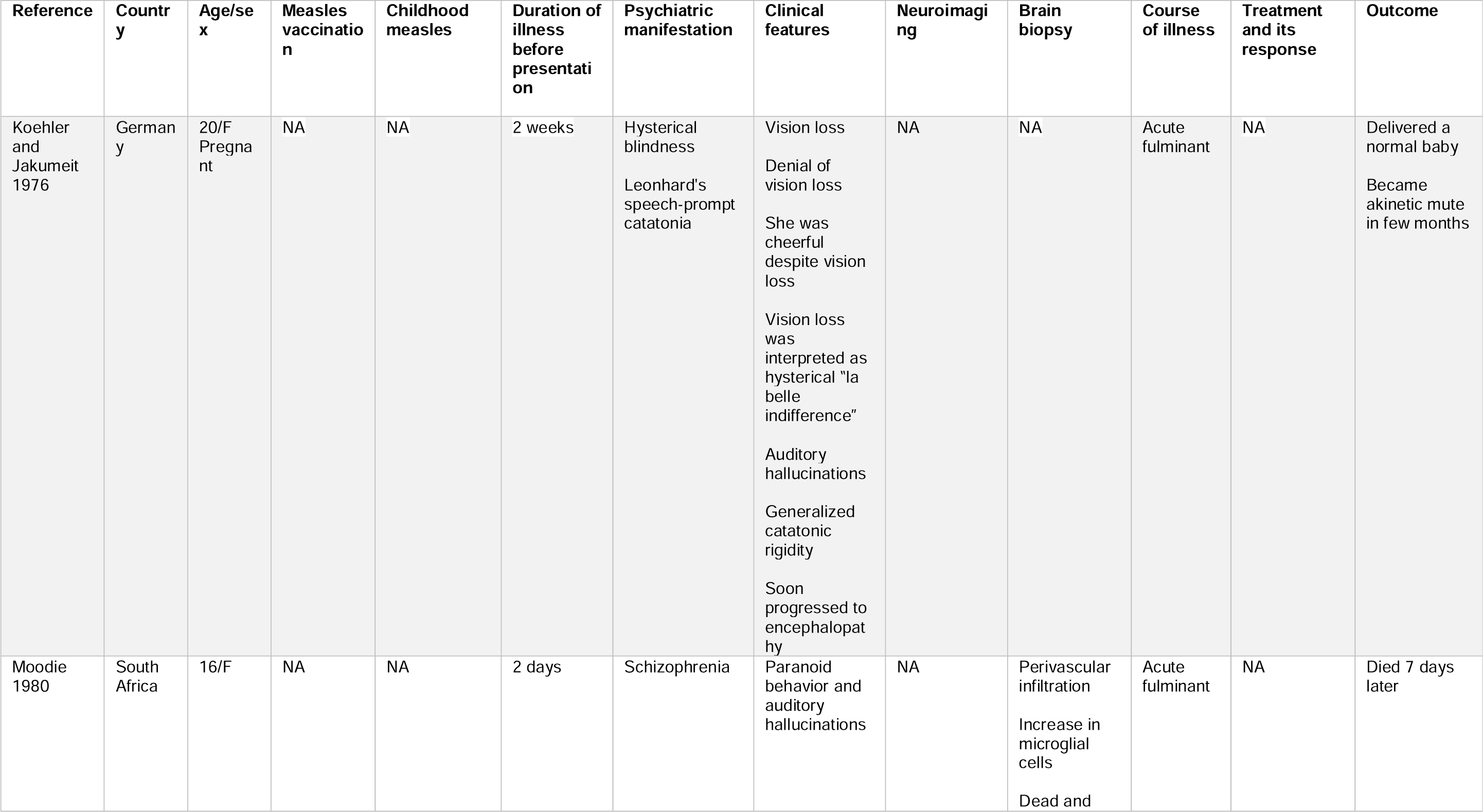
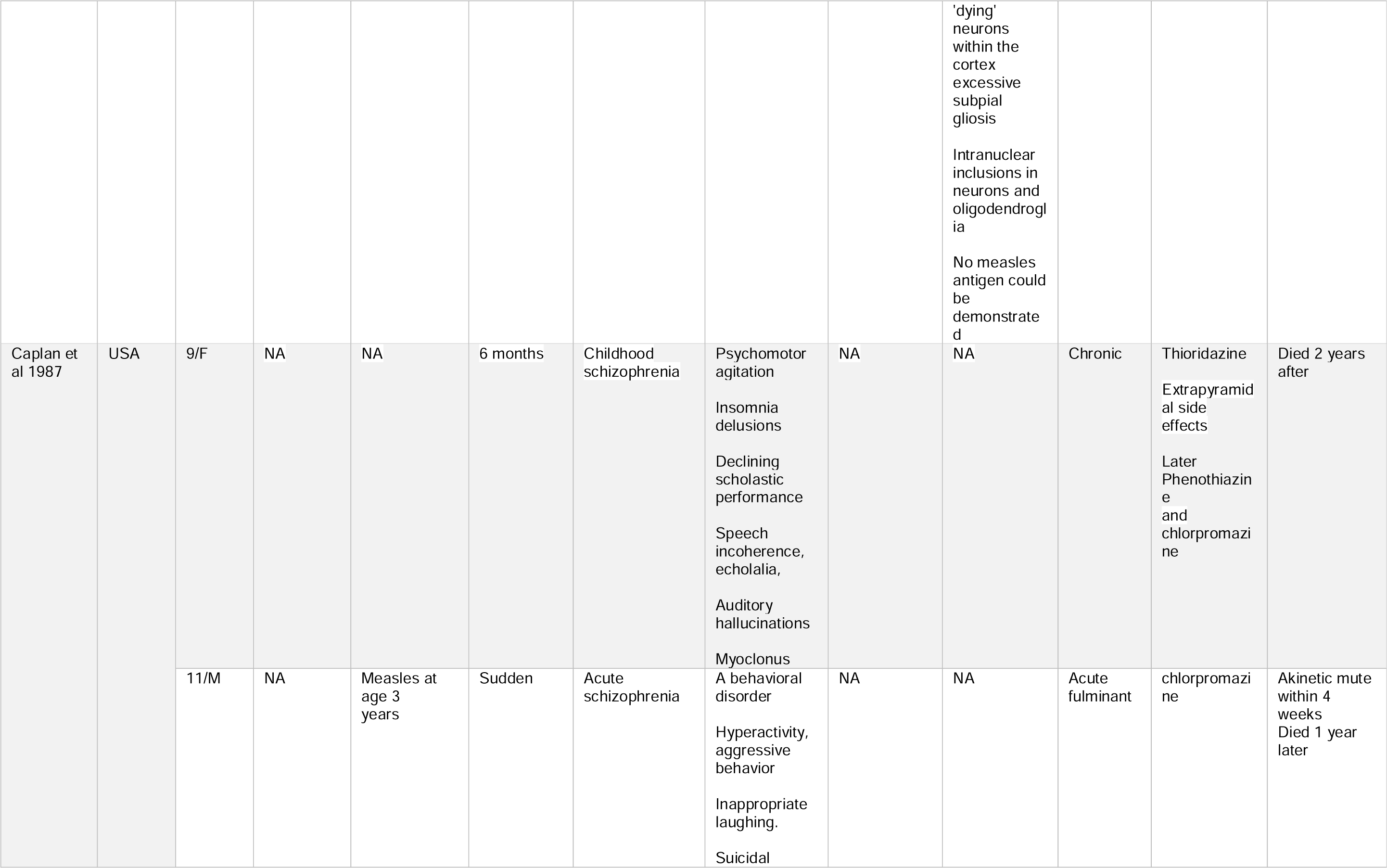
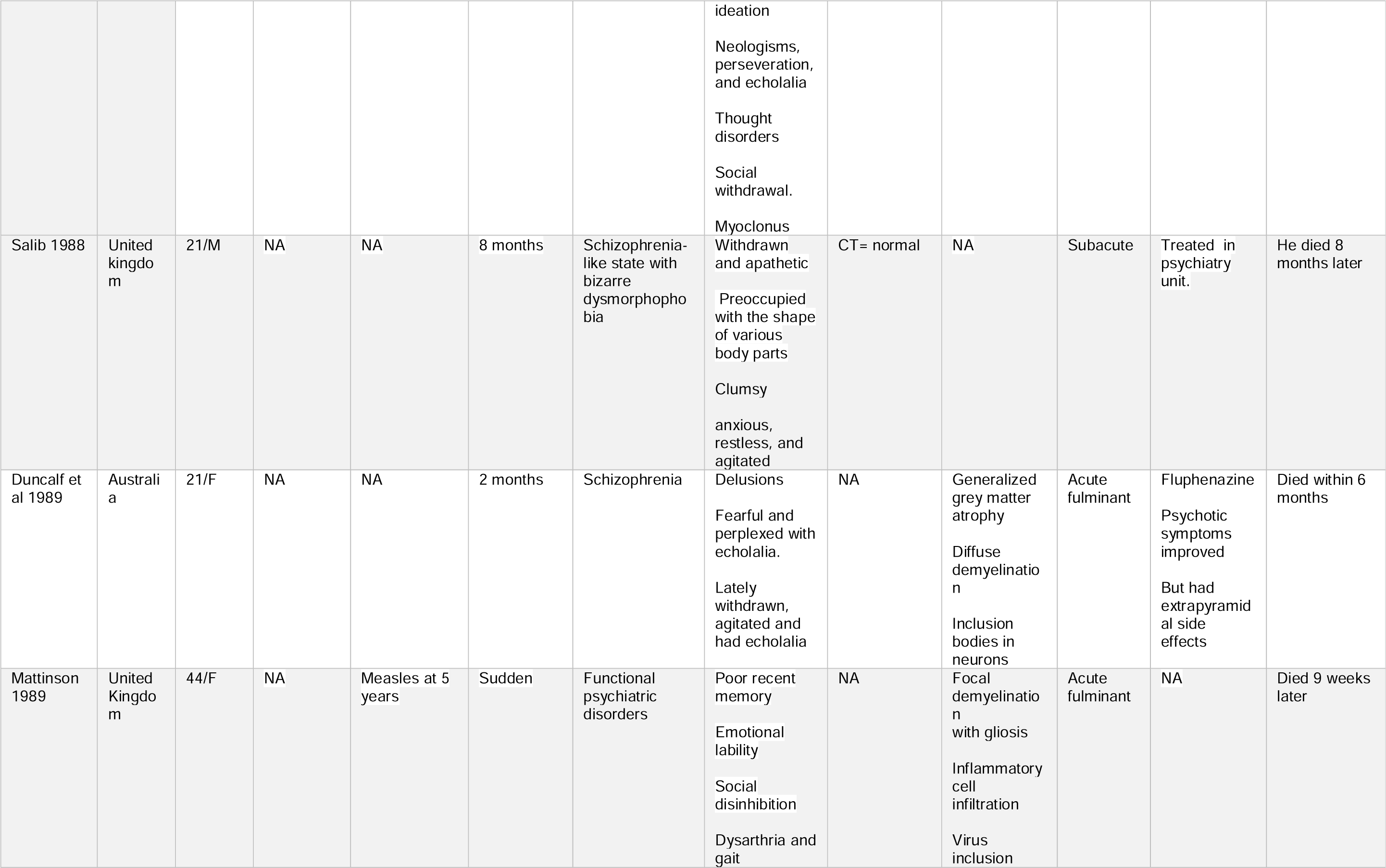
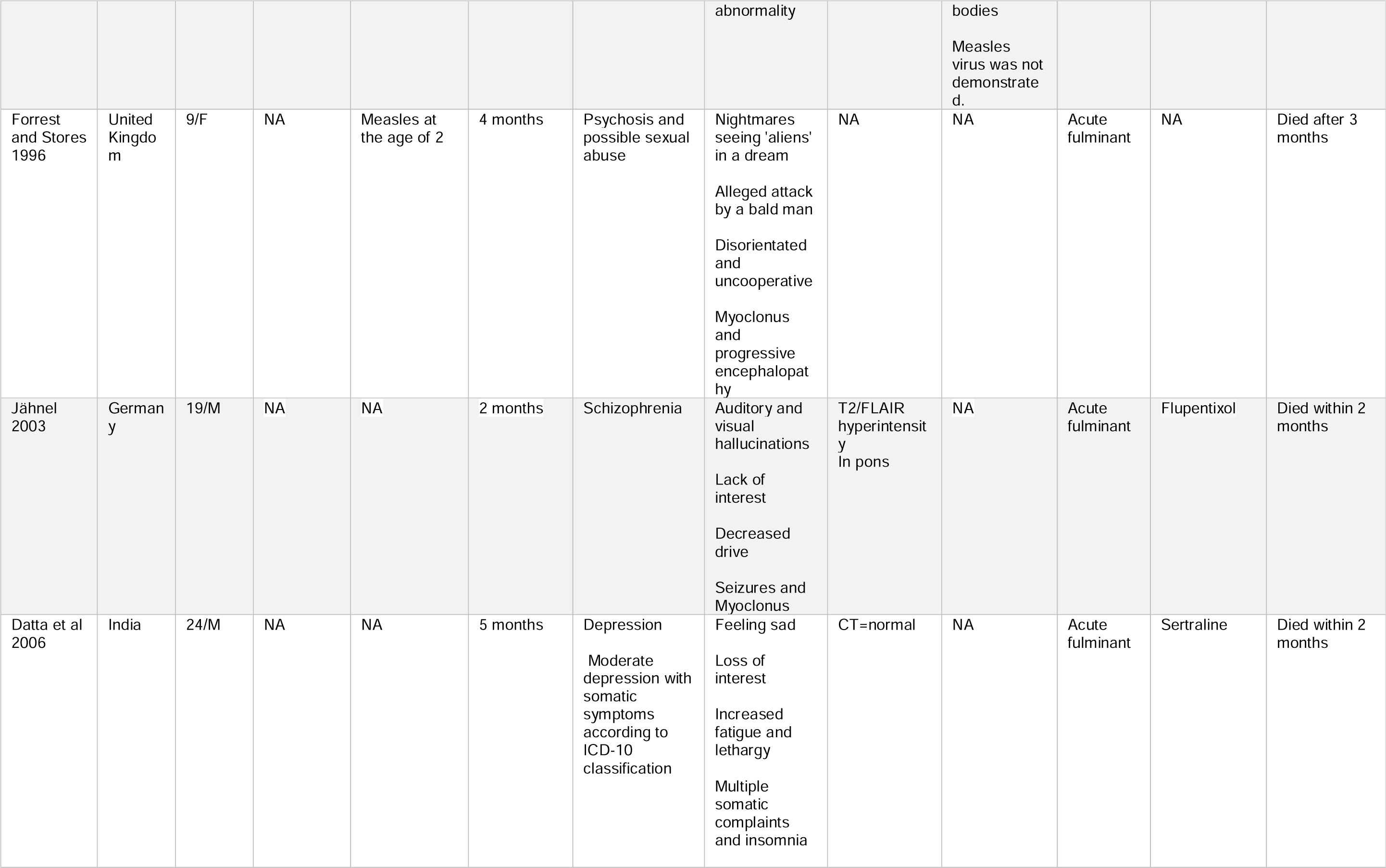
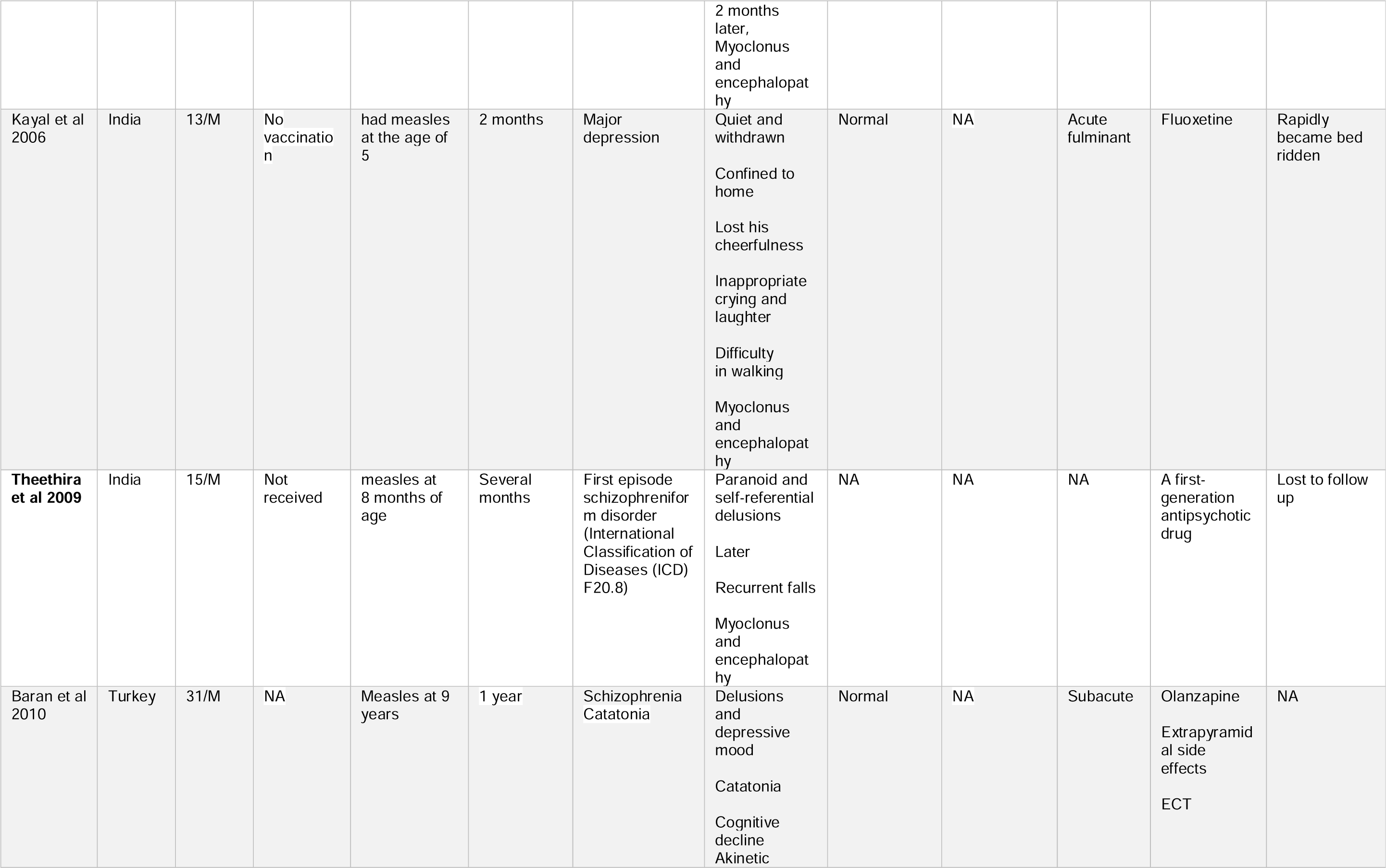
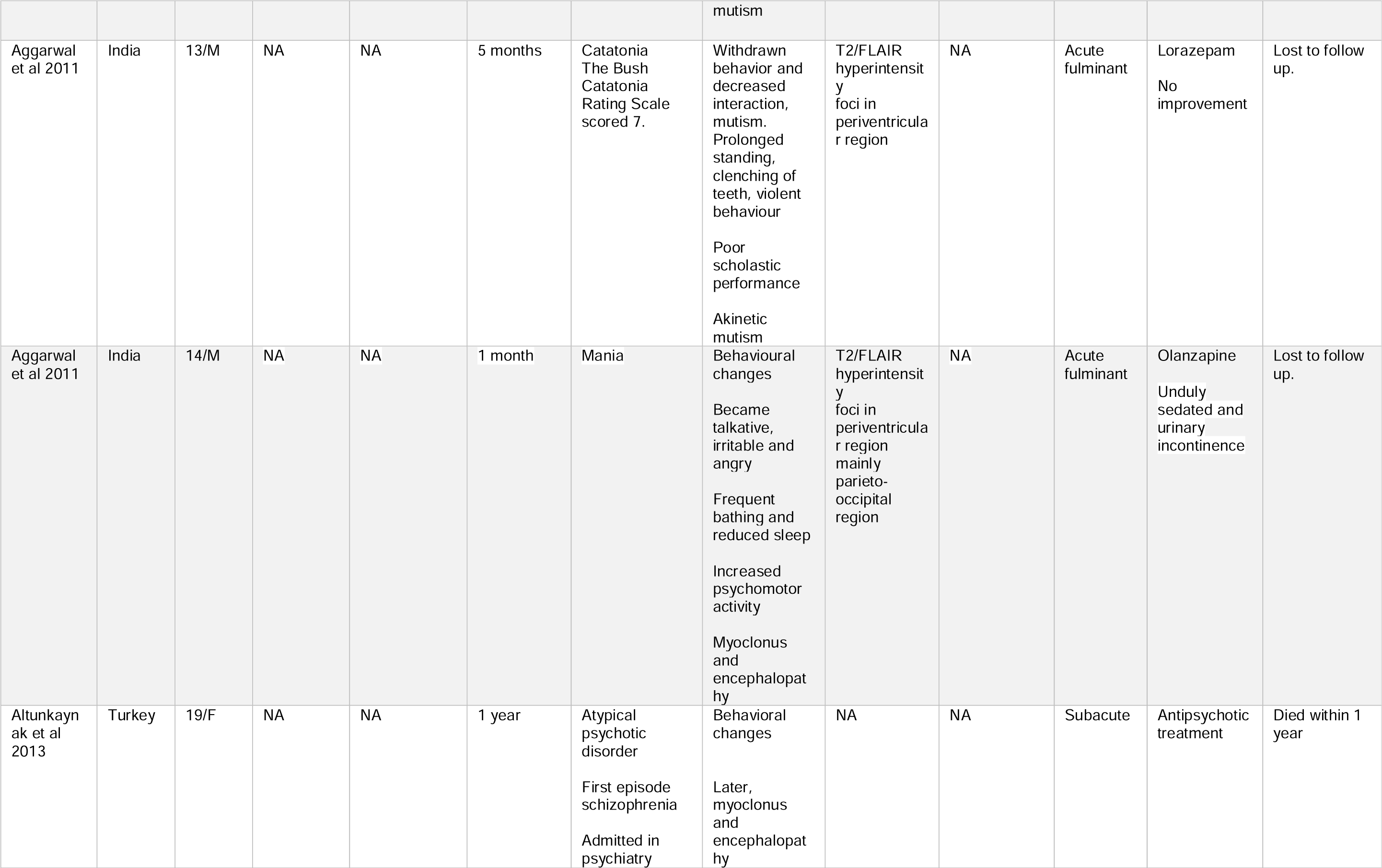
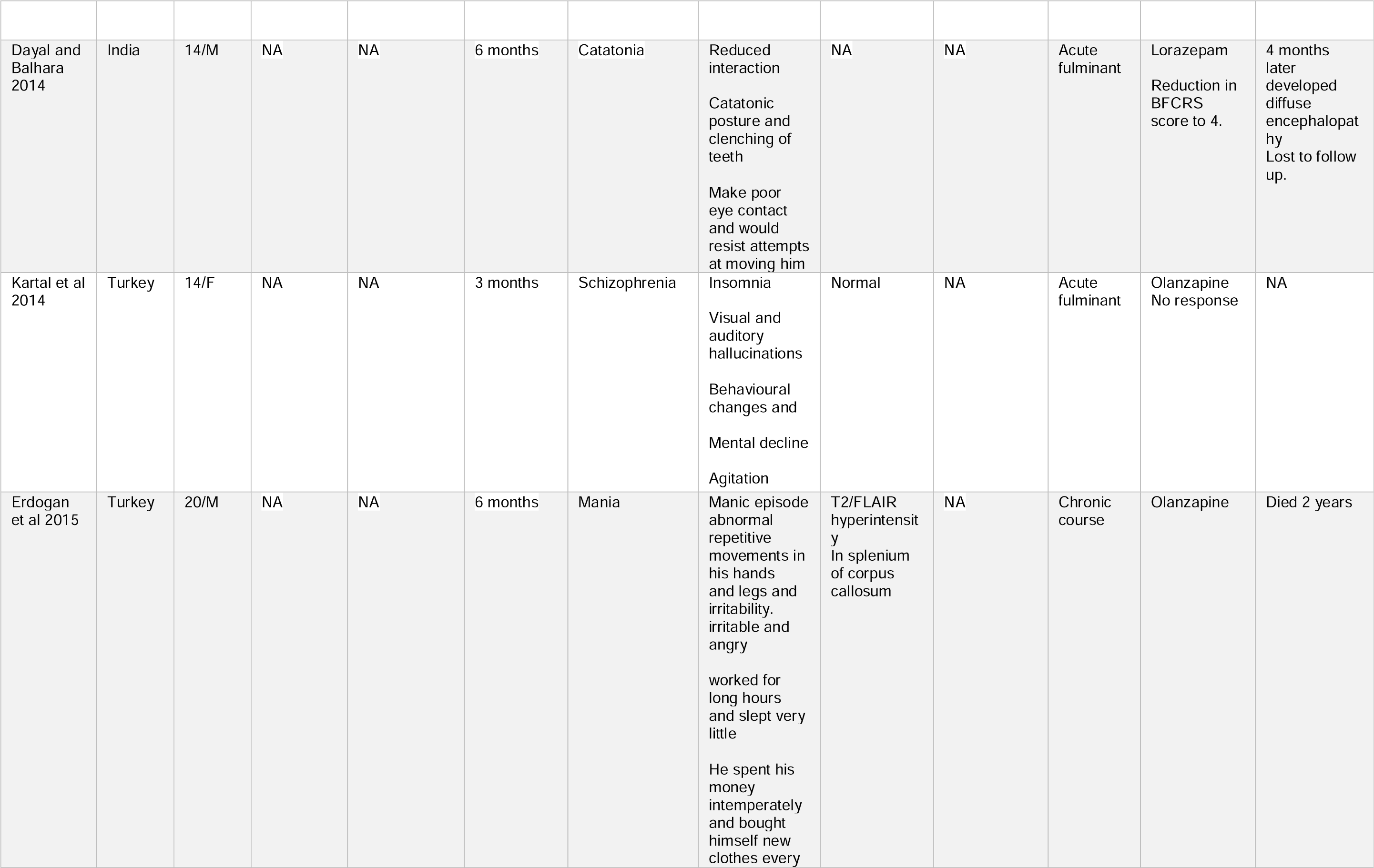
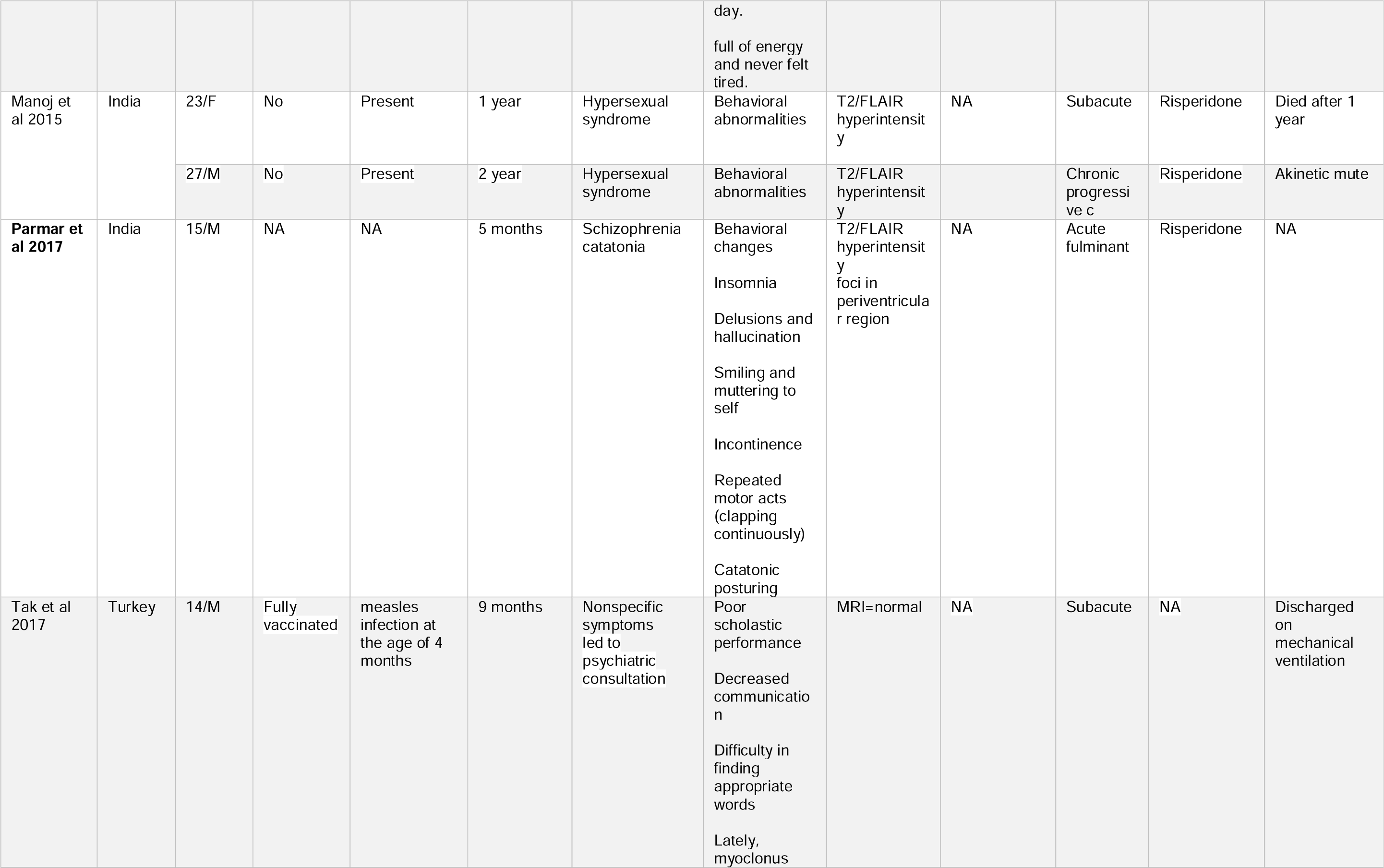
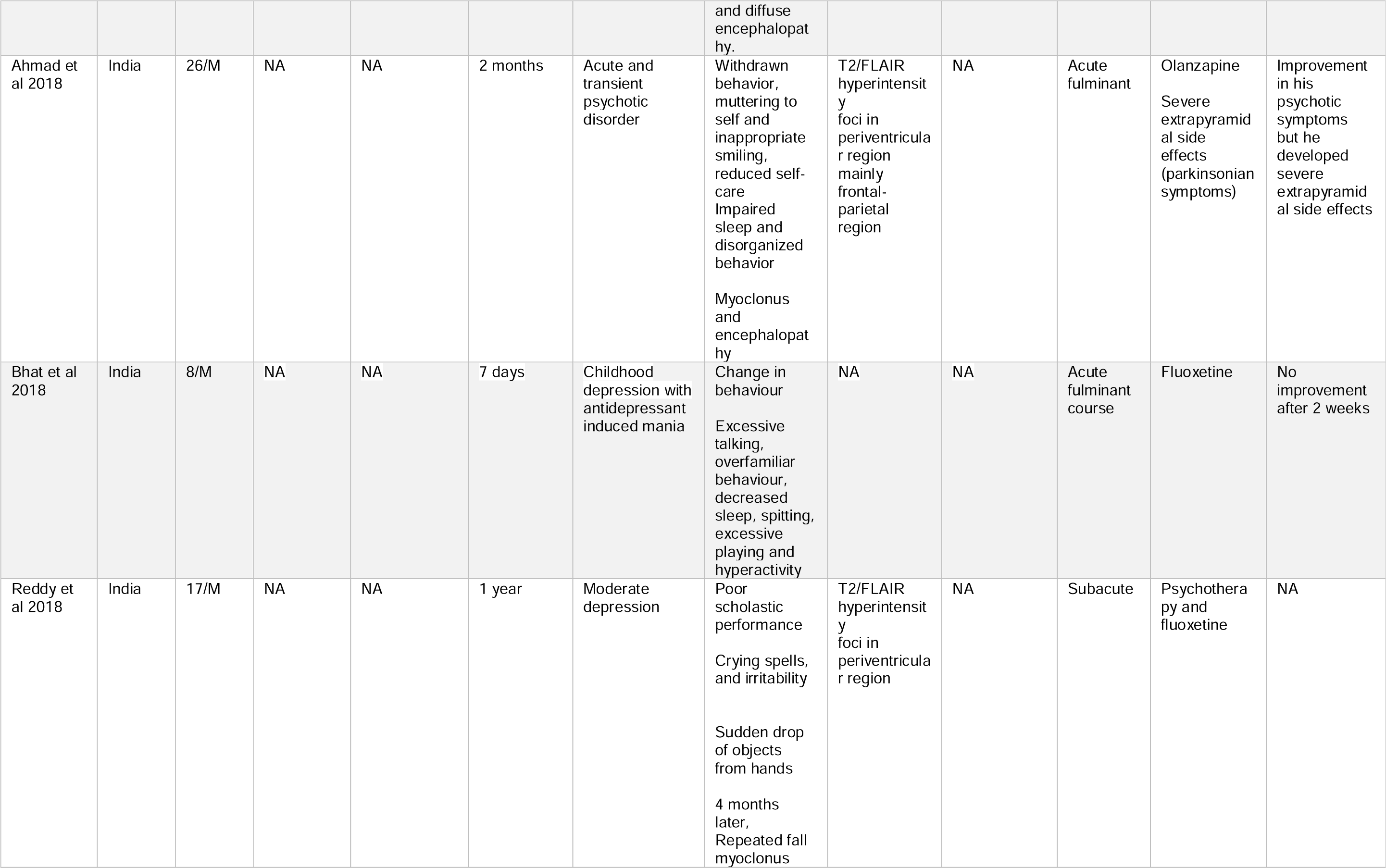
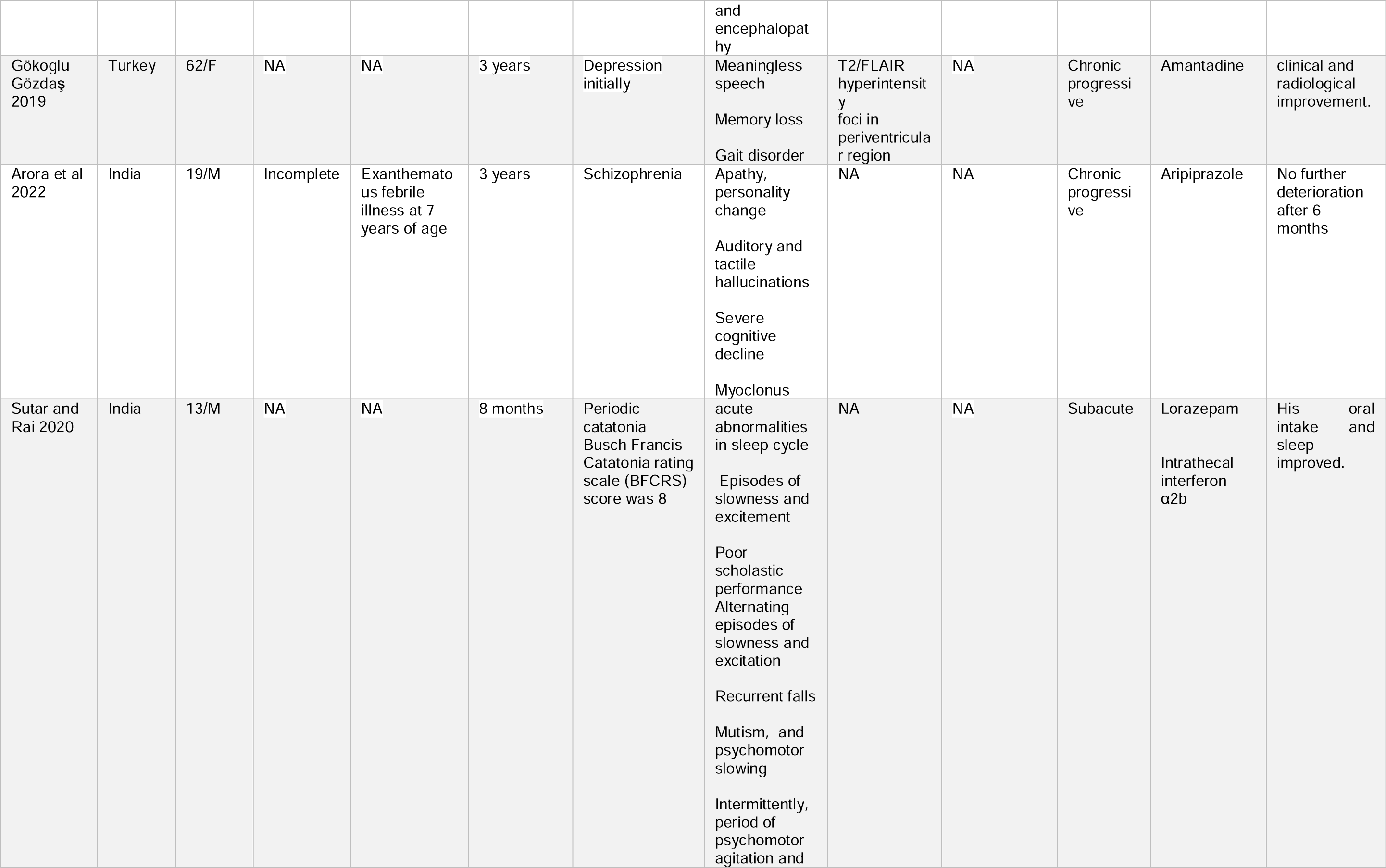
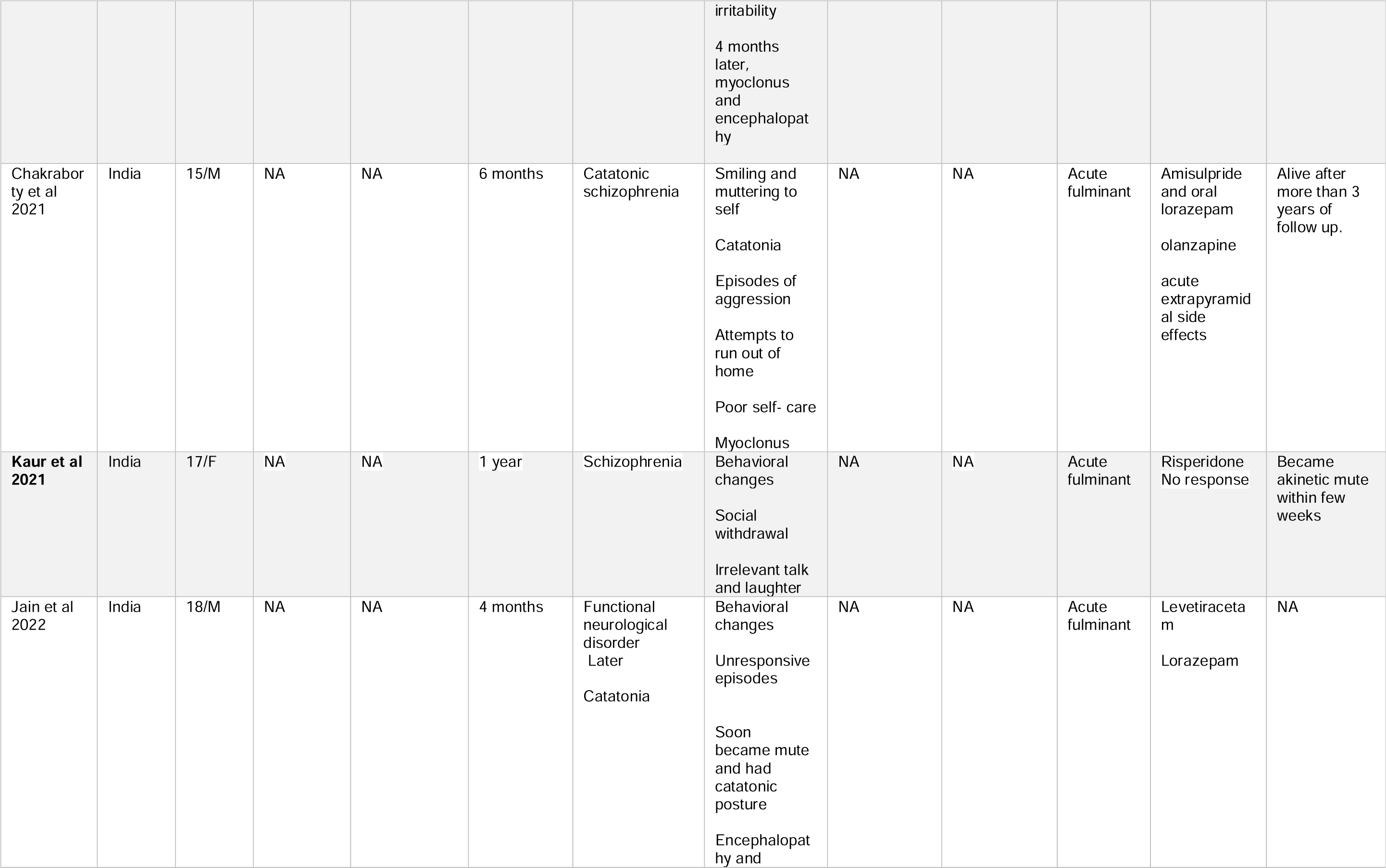
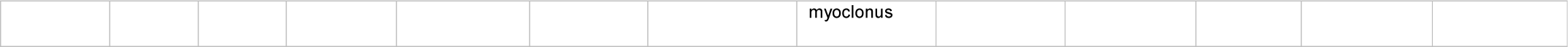
Epidemiological, clinical features, neuroimaging findings, histopathological features and outcome of patients of SSPE with psychiatric manifestations.

**Table-3:** Summary of epidemiological, clinical features, neuroimaging findings, histopathological features and outcome of patients of SSPE with psychiatric manifestations (n=32)

|  |  |
| --- | --- |
| Age (in years) | Mean= 19.4<br>Median=17 |
|  | Mode= 14<br>Range= 8-62<br>Interquartile Range= 7 |
| Sex | Female= 11 (34.3%)<br>Male= 21 (65.6%) |
| Measles vaccination | Yes= 2 (6.2%)<br>No/NA= 30 (93.7%) |
| Childhood measles | Yes= 10 (31.2%)<br>No or NA= 22 (68.8%) |
| Geographical areas of reported cases (total 30 reports) | India= 16 (53.3%)<br>Europe, except Turkey = 5 (16.7%)<br>Turkey= 6 (20%)<br>USA = 1 (3.3%)<br>Australia= 1 (3.3%)<br>Africa= 1 (3.3%) |
| course of the disease | Acute fulminant= 19 (59.3%)<br>Subacute= 7(21.9%)<br>Chronic =5 (16.6%)<br>NA=1 (3.1%) |
| Dominant psychiatric illness | Catatonia=3 (9.4%)<br>Depression= 5 (16.6%)<br>Functional neurological disorder/ Hysterical= 3 (9.4%)<br>Hypersexuality=2 (6.3%)<br>Mania= 2(6.3%)<br>Schizophrenia/ ill-defined psychotic disorders=16 (50%)<br>Non-specific= 1(3.1%) |
| Neuroimaging | NA= 16 (50%)<br>Normal= 6 (18.8%)<br>T2/FLAIR periventricular hyperintensity= 8 (25%)<br>Pontine hyperintensity= 1 (3.1%) |
|  | Splenium of the corpus callosum<br>hyperintensity =1 (3.1%) |
| Brain biopsy | NA=29 (90.6%)<br>Done=3 (9.4%) |

NA= not available; FLAIR= Fluid attenuated inversion recovery

A supplemental Word file contains the critical appraisal report of each included case, according to the Joanna Briggs Institute (JBI) checklist. (Supplementary file-2). In An 8-item scale, the majority of cases (26/32) complied with all the points. Only 6 cases faltered on the point “post-intervention clinical condition”.

The mean age of SSPE patients was 17.9 years (median 15 years and range 2-62 years). Men outnumbered the females (21: 11). Majority (94%) of SSPE patients with psychiatric presentations either did not have childhood measles vaccination, or a reliable vaccination history was not retrievable. In one-third of instances, childhood measles infection was reported by relatives. The majority of reports (73.3%) on SSPE patients with psychiatric presentations were from Turkey and India. After the year 2005, all such reports were excluded from these two countries only. (Table-3)

Schizophrenia, catatonia, and poorly characterized psychotic illnesses were the three most common psychiatric presentations (60%) in SSPE cases. Affective disorders, mania, and depression were reported among 23% (7/32) cases. In 10% of cases, the initial clinical presentation of SSPE was considered functional/ hysterical. In approximately 81% (26/32) cases the course of SSPE was rapidly progressive (either acute fulminant or subacute). (Table-3)

Neuroimaging abnormalities were detected in 63% (10/16) of patients, periventricular T2/FLAIR MR hyperintensities were the most frequently noted abnormalities. Brain biopsy findings were noted in 3 patients. Focal demyelination with gliosis, inflammatory cell infiltration, and the presence of inclusion bodies in neurons and oligodendroglia were dominant histopathological findings. (Table-3)

Treatment with antipsychotic drugs had poor or no response. Out of 17 patients who received antipsychotic drugs 6 patients noted severe extrapyramidal adverse effects, that further deteriorated the clinical condition of the patients. (Table-3)

## Discussion

In this systematic review, we noted that many patients with SSPE were mistakenly referred to psychiatric care because of their psychiatric presentations. These patients frequently received diagnoses of affective disorders, including mania and depression, as well as psychotic diseases like schizophrenia, catatonia, and poorly defined psychotic illnesses. Treatment with antipsychotic drugs often failed, and severe extrapyramidal adverse effects were common. In the early stages, many children were misdiagnosed as malingering or hysterical.

Our review found that schizophrenia was the most common psychiatric diagnosis for many cases of SSPE. Schizophrenia is a mental disorder characterized by delusions, hallucinations, disorganized thinking, and other symptoms. Anti-N-methyl-d-aspartate (NMDA) receptor encephalitis is another brain disorder that can cause similar symptoms. It is often misdiagnosed as a psychiatric disorder, just as SSPE was in our review. NMDA receptor encephalitis is characterized by psychotic and affective symptoms, catatonic signs, and mental decline. It is also often associated with seizures, abnormal movements, and a fluctuating course. Fulminant SSPE can mimic NMDA receptor encephalitis, making it difficult to distinguish between the two conditions..**^36-38^**

The pathophysiology of underlying schizophrenia in SSPE is difficult to elucidate. The psychotic symptoms may result from the progressive destruction of brain tissue caused by chronic measles virus infection. One report indicated that individuals with persistent schizophrenia had significantly higher blood antimeasles antibody titers than those without a history of a psychiatric disorder.**^39^** Many studies have shown that schizophrenia is associated with lower brain volume particularly of gray matter and cortical thickness, increasing gray matter loss, and abnormal gyral patterns.**^40^** Research has shown that there are structural changes in the brains of individuals with schizophrenia, and similar changes have been observed in the brains of individuals with SSPE. These changes include a reduction in brain volume, particularly in the frontal lobes, temporal lobes, and hippocampus, which are regions of the brain involved in cognitive and emotional processing. In patients with schizophrenia, neuropathological examinations and brain imaging studies suggest a variety of abnormalities in many brain regions, including the cerebral cortex, hippocampus, thalamus, and amygdala.**^41^** Extrapyramidal reactions following treatment with antipsychotic medications are common and frequently lead to rapid deterioration. (Figure-2)

**Figure-2.**
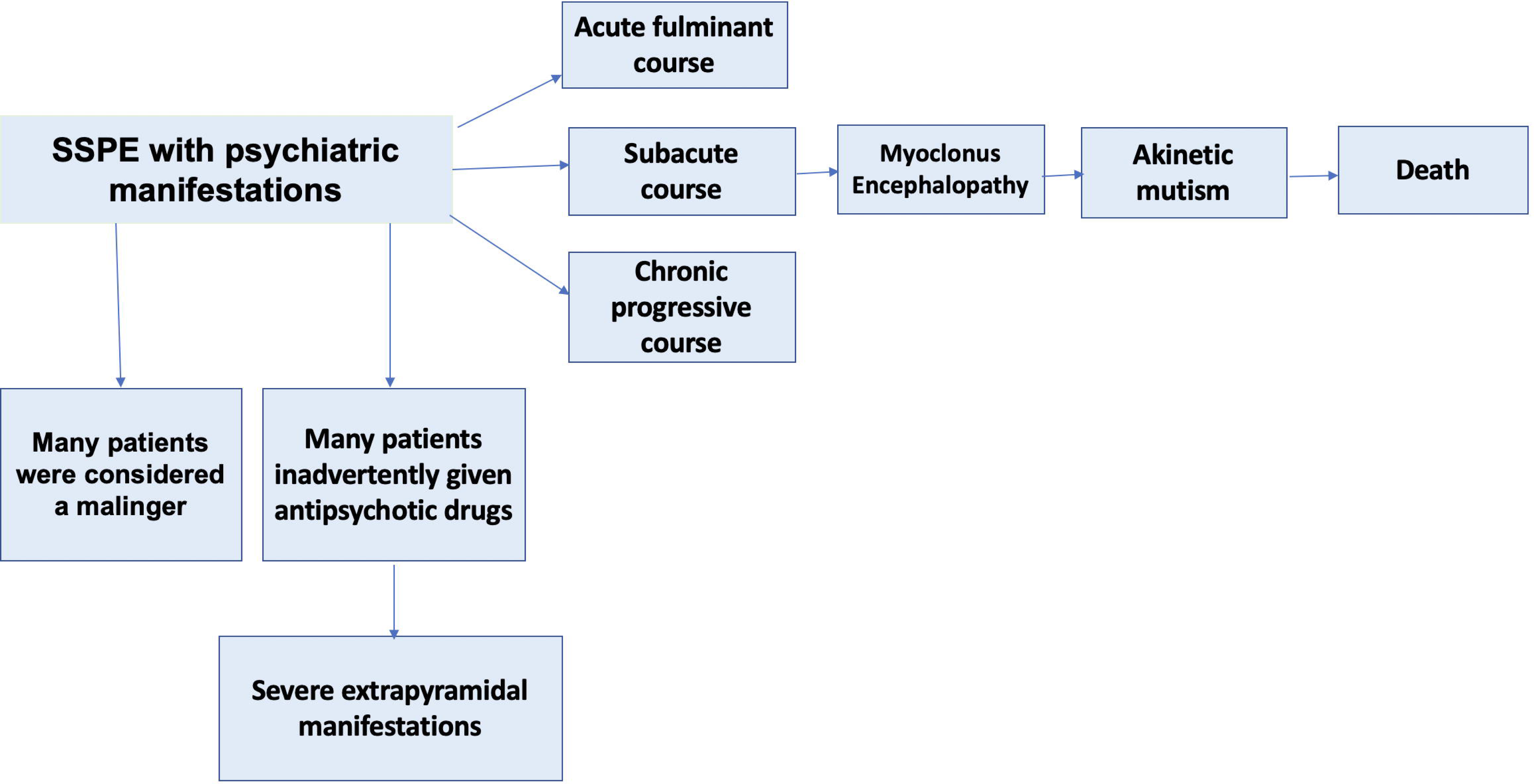
Flow diagram depicts progression of course of disease in SSPE patients with psychiatric manifestations.

Catatonia is another frequent psychiatric manifestation in patients with SSPE. The pathogenesis of catatonia in SSPE is also not fully understood. Catatonia is believed to result from dysfunction of the frontostriatal network in the brain, which regulates motor and behavioral functions. The SSPE virus also targets the basal ganglia, which are a part of the frontostriatal circuitry. Damage to the basal ganglia can disrupt the normal functioning of this circuitry, leading to motor and behavioral abnormalities characteristic of catatonia. The measles virus targets the limbic system, which is responsible for emotional regulation. Damage to the limbic system may lead to dysfunction of the neural circuits involved in mood regulation, leading to depression. In addition to direct damage to brain tissue, alterations in neurotransmitter and immune system function are possible mechanisms involved in the pathogenesis of depression in SSPE.**^42-44^**

Many times, particularly in the early stages of the disease, SSPE is inadvertently considered “malingering”. “Malingering” refers to the intentional feigning or exaggeration of symptoms for external gain, such as to avoid school work, or to gain attention or sympathy.**^45,46^**

In conclusion, many patients with SSPE inadvertently end up in psychiatric care due to their psychiatric symptoms. Early psychiatric manifestations in SSPE are often subtle and the diagnosis of SSPE is easily missed. We suggest that if there is no satisfactory response to antipsychotic drugs or if there are early extrapyramidal reactions leading to further deterioration, the possibility of SSPE should be considered.

## Declarations

Financial or non-financial support for the review= None

Competing interests of review authors= None to declare

## Supporting information

Supplementary item-1

Supplementary item-2

## Data Availability

All data produced in the present work are contained in the manuscript.

