## Supplementary item-1 for "The spectrum of psychiatric manifestations in subacute sclerosing panencephalitis: a systematic review of published case reports and case series"

**Supplementary item-1: The Joanna Briggs Institute Critical Appraisal tool for case reports**

| **Reference** | **Demographic characteristics** | **Patient’s history** | **Current clinical condition** | **Diagnostic assessment** | **Intervention(s) or treatment** | **Post-intervention condition** | **Adverse events (harms)** | **Takeaway lessons** | **Overall appraisal** |
| --- | --- | --- | --- | --- | --- | --- | --- | --- | --- |
| **Aggarwal et al 2011** | **YES** | **YES** | **YES** | **YES** | **YES** | **YES** | **YES** | **YES** | **8** |
| **Aggarwal et al 2011** | **YES** | **YES** | **YES** | **YES** | **YES** | **YES** | **YES** | **YES** | **8** |
| **Ahmad et al 2018** | **YES** | **YES** | **YES** | **YES** | **YES** | **YES** | **YES** | **YES** | **8** |
| **Arora et al 2022** | **YES** | **YES** | **YES** | **YES** | **YES** | **YES** | **YES** | **YES** | **8** |
| **Baran et al 2010** | **YES** | **YES** | **YES** | **YES** | **YES** | **NO** | **YES** | **YES** | **7** |
| **Bhat et al 2018** | **YES** | **YES** | **YES** | **YES** | **YES** | **YES** | **YES** | **YES** | **8** |
| **Caplan et al 1987** | **YES** | **YES** | **YES** | **YES** | **YES** | **YES** | **YES** | **YES** | **8** |
|  | **YES** | **YES** | **YES** | **YES** | **YES** | **YES** | **YES** | **YES** | **8** |
| **Chakraborty et al 2021** | **YES** | **YES** | **YES** | **YES** | **YES** | **YES** | **YES** | **YES** | **8** |
| **Datta et al 2006** | **YES** | **YES** | **YES** | **YES** | **YES** | **YES** | **YES** | **YES** | **8** |
| **Dayal and Balhara 2014** | **YES** | **YES** | **YES** | **YES** | **YES** | **YES** | **YES** | **YES** | **8** |
| **Duncalf et al 1989** | **YES** | **YES** | **YES** | **YES** | **YES** | **YES** | **YES** | **YES** | **8** |
| **Erdogan et al 2015** | **YES** | **YES** | **YES** | **YES** | **YES** | **YES** | **YES** | **YES** | **8** |
| **Forrest and Stores 1996** | **YES** | **YES** | **YES** | **YES** | **YES** | **YES** | **YES** | **YES** | **8** |
| **Gökoglu Gözdaş 2019** | **YES** | **YES** | **YES** | **YES** | **YES** | **YES** | **YES** | **YES** | **8** |
| **Jähnel 2003** | **YES** | **YES** | **YES** | **YES** | **YES** | **YES** | **YES** | **YES** | **8** |
| **Jain et al 2021** | **YES** | **YES** | **YES** | **YES** | **YES** | **NO** | **YES** | **YES** | **7** |
| **Kartal et al 2014** | **YES** | **YES** | **YES** | **YES** | **YES** | **NO** | **YES** | **YES** | **7** |
| **Kaur et al 2021** | **YES** | **YES** | **YES** | **YES** | **YES** | **YES** | **YES** | **YES** | **8** |
| **Kayal et al** | **YES** | **YES** | **YES** | **YES** | **YES** | **YES** | **YES** | **YES** | **8** |
| **Koehler and Jakumeit 1976** | **YES** | **YES** | **YES** | **YES** | **YES** | **YES** | **YES** | **YES** | **8** |
| **Manoj et al 2015** | **YES** | **YES** | **YES** | **YES** | **YES** | **YES** | **YES** | **YES** | **8** |
|  | **YES** | **YES** | **YES** | **YES** | **YES** | **YES** | **YES** | **YES** | **8** |
| **Mattinson 1989** | **YES** | **YES** | **YES** | **YES** | **YES** | **YES** | **YES** | **YES** | **8** |
| **Moodie 1980** | **YES** | **YES** | **YES** | **YES** | **YES** | **YES** | **YES** | **YES** | **8** |
| **Panensefalit 2013** | **YES** | **YES** | **YES** | **YES** | **YES** | **YES** | **YES** | **YES** | **8** |
| **Parmar et al 2017** | **YES** | **YES** | **YES** | **YES** | **YES** | **NO** | **YES** | **YES** | **7** |
| **Reddy et al 2018** | **YES** | **YES** | **YES** | **YES** | **YES** | **NO** | **YES** | **YES** | **7** |
| **Salib 1988** | **YES** | **YES** | **YES** | **YES** | **YES** | **YES** | **YES** | **YES** | **8** |
| **Sutar and Rai 2020** | **YES** | **YES** | **YES** | **YES** | **YES** | **YES** | **YES** | **YES** | **8** |
| **Tak et al 2017** | **YES** | **YES** | **YES** | **YES** | **YES** | **YES** | **YES** | **YES** | **8** |
| **Theethira et al 2009** | **YES** | **YES** | **YES** | **YES** | **YES** | **NO** | **YES** | **YES** | **7** |

**The Joanna Briggs Institute Critical**

**Appraisal tool for case reports consists of following 8 items**

1. **Were patient’s demographic characteristics clearly described?**
2. **Was the patient’s history clearly described and presented as a timeline?**
3. **Was the current clinical condition of the patient on presentation clearly described?**
4. **Were diagnostic tests or assessment methods and the results clearly described?**
5. **Was the intervention(s) or treatment procedure(s) clearly described?**
6. **Was the post-intervention clinical condition clearly described?**
7. **Were adverse events (harms) or unanticipated events identified and described?**
8. **Does the case report provide takeaway lessons?**
